# The Impact of Instructions on Individual Prioritization Strategies in a Dual-Task Paradigm for Listening Effort

**DOI:** 10.1101/2024.06.26.24309528

**Authors:** Katrien Kestens, Emma Lepla, Flore Vandoorne, Dorien Ceuleers, Louise Van Goylen, Hannah Keppler

**Author notes:** All correspondence should be addressed to: Katrien Kestens, Faculty of Medicine and Health Sciences, Department of Rehabilitation Sciences, Corneel Heymanslaan 10 (2P1), Ghent University, 9000 Ghent, Belgium. **Declaration of Interest** None to report. **Data availability statement** The data that support the findings of this study are available from the corresponding author, KK, upon reasonable request.

## Abstract

**Introduction:** This study examined the impact of instructions on the prioritization strategy employed by individuals during a listening effort dual-task paradigm.

**Methods:** The dual-task paradigm consisted of a primary speech understanding task in different listening conditions and a secondary visual memory task, both performed separately (baseline) and simultaneously (dual-task). Twenty-three normal-hearing participants (mean age: 36.8 years; 14 females) were directed to prioritize the primary speech understanding task in the dual-task condition, whereas another twenty-three (matched for age, gender, and education level) received no specific instructions regarding task priority. Both groups performed the dual-task paradigm twice (mean interval: 14.8 days). Patterns of dual-task interference were assessed by plotting the dual-task effect of the primary and secondary task against each other. Fisher’s exact tests were used to assess whether there was an association between interference patterns and group (non-prioritizing and prioritizing) across all listening conditions and test sessions.

**Results:** No statistically significant association was found between the pattern of dual-task interference and the group to which the participants belong for any of the listening conditions and test sessions. Descriptive analysis revealed no consistent strategy use within individuals across listening conditions and test sessions, suggesting a lack of a uniform approach regardless of the given instructions.

**Conclusion:** Providing prioritization instructions was insufficient to ensure that an individual will mainly focus on the primary task and consistently adhere to this strategy across listening conditions and test sessions. These results raised reservations about the current usage of dual-task paradigms for listening effort.

## 1. INTRODUCTION

Listening effort – defined as *“the deliberate allocation of mental resources to overcome obstacles in goal pursuit during a [listening] task”* (Pichora-Fuller et al., 2016, p. 10) – is one of the main complaints of individuals with hearing loss and plays a vital role in listeners’ communication and quality of life (Cañete et al., 2023; Pang et al., 2019). One commonly reported approach to measure listening effort is a dual-task paradigm (e.g., Degeest et al., 2022; Gagne et al., 2017; Kuchinsky et al., 2024). In the context of listening effort, a dual-task paradigm evaluates an individual’s ability to understand speech (primary task) while concurrently performing another task (secondary task). Both tasks are performed separately and concurrently, denoted as baseline and dual-task condition, respectively.

Dual-task paradigms are based on Kahneman’s theory that the available resources to perform a task are limited in capacity (Kahneman, 1973). When the attentional and cognitive resources required for performing the primary and secondary tasks concurrently are lower than the total available resources, both tasks can be handled effectively. However, if the required resources to simultaneously perform the primary and secondary task exceed the listeners’ maximum cognitive capacity, the processing system will prioritize one of the tasks under the dual-task condition. In the context of listening effort, it is generally instructed to prioritize the primary speech understanding task (Gagne et al., 2017). One might assume that providing this prioritization instruction will result in stable scores on the primary task across both baseline and dual-task conditions. As a result, the change in secondary task performance from the baseline condition to the dual-task condition serves as a measure of listening effort (Gagne et al., 2017). In dual-task paradigms used to measure listening effort, the primary task is presented under various listening conditions, with changes in secondary task performance reflecting the increased difficulty of the primary task. Specifically, as the primary task becomes more challenging, individuals are expected to reallocate their cognitive resources to maintain optimal performance on the primary task, leading to a decline in secondary task performance. This change is typically referred to as dual-task cost in hearing research, highlighting the negative impact of an increasingly demanding primary task on secondary task performance (Gagné et al., 2017).

Intriguingly, a recent study challenged the assumption that prioritization instructions consistently lead to stable scores on the primary task across both baseline and dual-task conditions (Ceuleers et al., 2024). The study included 31 normal-hearing adults, 31 hearing aid users, and 31 cochlear implant users, all matched for age, gender, and educational level. Participants completed a dual-task paradigm consisting of a primary speech understanding task in three listening conditions of varying difficulty and a secondary visual memory task. Despite clear prioritization instructions, some participants failed to maintain consistent primary task scores, while others demonstrated improved secondary task performance under the dual-task condition compared to the baseline, particularly as the listening condition became more challenging. Notably, this phenomenon was observed across all three participant groups. This finding contradicts the theoretical assumption that increased difficulty in the primary task would invariably lead to declines in secondary task performance, even when prioritization instructions are provided. This suggests that we cannot assume participants will always strictly follow instructions and that they might unconsciously develop their own strategies when their maximum cognitive capacity is exceeded (Choi et al., 2008).

Consequently, it was concluded that the theoretical framework cannot be universally applied at the individual level and proposed an alternative approach for a more comprehensive interpretation of a listening effort dual-task paradigm (Ceuleers et al., 2024). This new approach, although grounded in earlier cognitive-motor dual-task research (Plummer & Eskes, 2015) is especially innovative in the field of hearing research, as it has never been applied in this context before. However, previous authors have suggested it as a potential metric for measuring listening effort (Degeest et al., 2021; Gagne et al., 2017).

In this approach, the dual-task effect (DTE, i.e. the change in task performance from the baseline condition to the dual-task condition) of both the primary and secondary tasks are calculated separately using the formula DTE (%) = 100 × [(score in dual-task condition - score in baseline condition)/score in baseline condition]. Negative DTE values imply a decrease in performance in the dual-task relative to the single-task (i.e. dual-task cost, - DTE), whereas positive DTE values suggest an advancement in performance in the dual-task (i.e. dual-task benefit, + DTE) (Plummer & Eskes, 2015). By plotting the DTE of the primary and secondary tasks against each other different patterns of dual-task interference can be distinguished (Figure 1). These patterns offer valuable insights into attention allocation, and consequently, the individual strategies employed when performing a dual-task paradigm (Plummer & Eskes, 2015; Yogev-Seligmann et al., 2012). For example, a dual-task cost in the secondary task may occur with a reciprocal dual-task cost (i.e. mutual interference), no change (i.e. visual memory interference), or a dual-task benefit (i.e. speech priority trade off) in the primary task. All three scenarios imply that the attentional demands exceed the total capacity to maintain single-task-level performance but differ in the trade-off of attentional resources. For example in the visual memory interference scenario, the listener prioritized the primary task such that only the secondary task deteriorates while primary performance maintained. The speech priority trade off scenario suggests a trade-off in attentional resources, such that improved performance in the primary task occurred at a cost to the secondary task.

**Figure 1:**
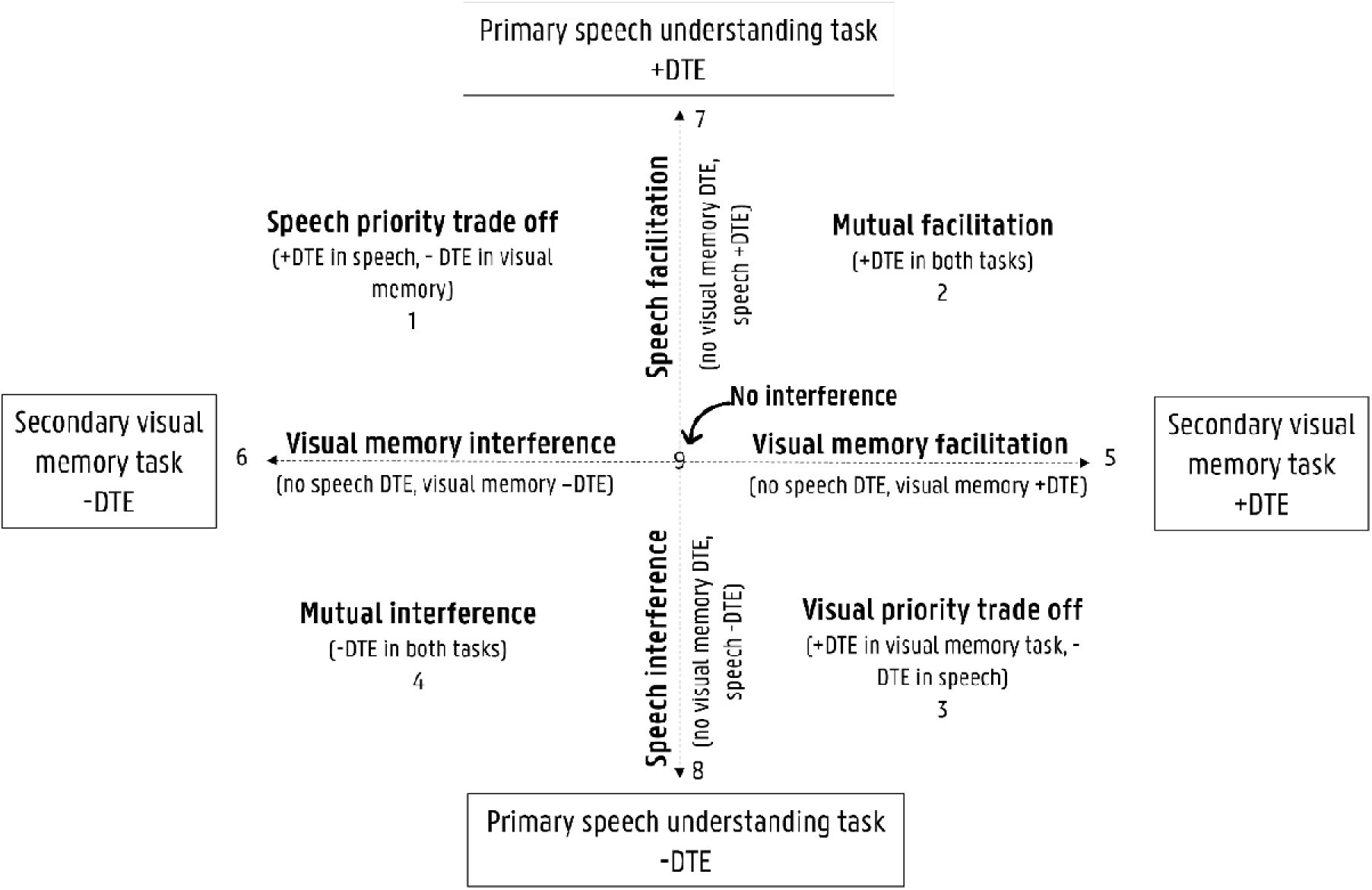
Framework of the dual-task interference patterns by plotting the dual-task effect (DTE) of the primary and secondary task against each other, based on Plummer and Eskes (2015).

Previous research with children demonstrated that instructions alone were ineffective in controlling listeners’ strategies when performing a dual-task paradigm. Specifically, instructions were insufficient to ensure that participants prioritized the primary speech understanding task in a listening effort dual-task paradigm (Choi et al., 2008). However, this phenomenon remains understudied in adults. To address this gap, the current study investigated the influence of instructions on adults’ prioritization strategies during a listening effort dual-task paradigm across various listening conditions and test sessions. We analyzed both group-level performance stability on the primary task (the standard approach) and individual patterns of dual-task interference (the new approach). Drawing on previous findings (Ceuleers et al., 2024; Choi et al., 2008), we hypothesized that task prioritization strategies are individually determined and may vary over time. Factors such as listener characteristics (e.g. age, hearing sensitivity, personality, motivation) and environmental conditions (e.g. background noise and reverberation) are known to influence how attentional and cognitive resources are allocated to understand speech (Kuchinsky et al., 2023; Peelle, 2018; Pichora-Fuller et al., 2016; Strand et al., 2018). Likewise, we anticipated substantial interindividual variability in prioritization strategies, regardless of the given prioritization instructions. Hence, we assumed that individual patterns of dual-task interference offer more accurate insights into task prioritization strategies than group-level performance stability.

Understanding how listeners execute a dual-task paradigm for listening effort provide a critical benchmark for future studies and potential clinical applications in managing listening challenges.

## 2. METHODS

The current study was approved by the local Ethics Committee (reference number ONZ-2022-0204) and was conducted in accordance with the Helsinki Declaration. Prior to participation, an informed consent was signed. No financial reward was given to the participants.

### 2.1 Participants

Normal-hearing adults between 18 and 69 years of age were recruited through convenience sampling between August and December 2022. All participants were native Flemish speakers who had completed a minimum of 12 years of education (equivalent to high school graduation). All participants had either normal or corrected-to-normal vision, as assessed by the Near Vision Snellen Eye Chart (Snellen, 1873). Exclusion criteria encompassed chronic tinnitus (≥3 months), self-reported excessive noise exposure within the past 72 hours, attention deficits, as well as learning, psychiatric, or (a history of) neurological disorders. For participants aged 60 years and older, the Montréal Cognitive Assessment (with a cutoff score of 26) was administered to screen for potential cognitive impairment (Nasreddine et al., 2005).

Bilateral normal middle-ear status was confirmed for all participants through otoscopy and 226 Hz tympanometry (TympStar Pro). Air-conduction pure-tone thresholds were obtained bilaterally through headphones for octave frequencies between 250 and 8000 Hz using the modified Hughson-Westlake method (Equinox 2.0 audiometer with TDH39 headphone, Interacoustics). Pure-tone audiometry, performed in a sound-treated booth, adhered to the International Organization for Standardization (ISO) 8253-1 guideline for ambient sound pressure levels, ensuring accurate testing ((ISO, 2010). Participants were included when air-conduction thresholds at each tested octave frequency were symmetrical and when the better-ear audiometric threshold averaged over 0.5, 1, 2, and 4 kHz was equal or below 20 dB HL (World Health Organization, 2021). Asymmetric hearing was identified as a difference of 15 dB or more between the right and left ears at three consecutive frequencies.

### 2.2 Dual-task paradigm

A dual-task paradigm, based on that reported in Degeest et al. (2015), was used. The primary and secondary task consisted respectively of a speech understanding and visual memory task, and were both performed separately (baseline condition) and simultaneously (dual-task condition).

#### 2.2.1 Baseline condition

The primary task was a speech understanding task presented in different listening conditions, varying in listening demand. Words of the *Brugge-Leuven-Utrecht* (BLU) list were used as speech stimuli. This BLU list is endorsed by the Belgian National Institute for Health and Disability Insurance in, among other things, the context of hearing aid reimbursement. Consequently, in Belgium, this word list is considered gold standard within daily audiological practice and was, therefore, used within the current study. The BLU list contains two-syllable words. All words were spoken by a native Flemish female speaker without coarticulation. A steady state noise was generated and spectrally shaped to reflect the long-term average speech spectrum of the speech material. Words and noise were digitally mixed to create different signal-to-noise ratios (ranging from +8 dB SNR to −4 dB SNR in steps of 4 dB) whereby a constant noise level (i.e. 65 dB SPL) was utilized to which the intensity level of the speech stimuli was altered. Before stimulus onset, 10 seconds noise followed by a one second attention pure-tone of 1000 Hz were played to adapt to the noise and to focus participants’ attention to stimulus presentation, respectively. In addition to these noise conditions, a quiet listening condition without background noise was used. Within each listening condition, three trials of five words were presented. The words and background noise were presented through two loudspeakers, positioned at 45° and 315° azimuth at a distance of 90 cm from the participant’s ear. After each trial, participants verbally repeated the words. A word scoring was used, leading to a maximum score of 15 per listening condition.

The secondary task was a visual memory task in which a raster (i.e. 12 separated squares) was shown on a white standard computer screen (see example in Degeest et al. (2015)). Participants were seated on a chair and sat 70 cm in front of the computer, positioned at eye height. Within the raster, series of five identical blue filled circles appeared successively for 1 s each. Participants had to memorize the squares in which the circles appeared and had to indicate the position on a score form. It was obligated to indicate five squares, even if guessing was necessary. For each square correctly designated, one point was assigned. Five trials of five circles were presented. To determine the baseline score for the secondary task, only the score of the fourth and fifth trials were used in order to control for learning effects (Degeest et al., 2015). Eventually, this score on ten was multiplied by 1.5, to reach a score on 15, making it comparable with the primary scores.

#### 2.2.2 Dual-task condition

In the dual-task condition, the primary speech understanding task and secondary visual memory task were performed simultaneously. Participants underwent three trials where sequences of five words were presented alongside the appearance of five circles. This occurred under each of the five fixed listening conditions (quiet, SNR +8 dB, SNR +4 dB, SNR 0 dB, SNR −4 dB SNR). To minimize any potential conditioning effect, the auditory presentation of a word was not perfectly synchronized with the visual appearance of a circle, and could begin slightly before or after the appearance of the circle. Participants had to verbally repeat the words, after which they assigned the position of the squares on the score form. For each correctly repeated word and correctly designated square, one point was counted for the primary and secondary task respectively, leading to a maximum score of 15 per listening condition for both tasks.

#### 2.2.3 Dual-task interference

Each participant’s dual-task interference was measured by calculating the DTE for both the primary and secondary task separately (DTE = 100 × [score in dual-task condition - score in baseline condition]/score in baseline condition) (Gagne et al., 2017; Plummer & Eskes, 2015). By plotting the DTE of the primary and secondary task against each other (Figure 1), nine distinct patterns of dual-task interference can be identified: (1) speech priority trade off (dual-task benefit in the speech understanding task, dual-task cost in the visual memory task), (2) mutual facilitation (dual-task benefit in both tasks), (3) visual priority trade off (dual-task benefit in the visual memory task, dual-task cost in the speech understanding task), (4) mutual interference (dual-task cost in both tasks), (5) visual memory facilitation (no change in the speech understanding task, dual-task benefit in the visual memory task), (6) visual memory interference (no change in the speech understanding task, dual-task cost in the visual memory task), (7) speech facilitation (no change in the visual memory task, dual-task benefit in the speech understanding task), (8) speech interference (no change in the visual memory task, dual-task cost in the speech understanding task), and (9) no interference (no change in both tasks) (Plummer & Eskes, 2015).

### 2.3 Test procedure

The dual-task paradigm was carried out in a quiet, non-reverberant room illuminated with daylight and standard room lighting. At the beginning of each test, a practice trial was performed to verify whether the task was understood. If not, additional instructions were given until the participants fully understood the task.

To assess the effect of instructions on the prioritization strategy, participants were divided into two groups: the prioritizing and non-prioritizing group. The instruction provided during dual-task testing varied between the groups. In the prioritizing group, participants were directed to prioritize the primary speech understanding task (Gagne et al., 2017). Conversely, the non-prioritizing group received no specific instructions regarding task priority. The initial participants were randomly assigned to either the prioritizing or non-prioritizing group, after which matching participants were selected for the other group. To account for influencing factors, one-on-one matching was conducted based on age (±2 years), gender, and educational level (≤ 12 years of education or > 12 years of education). The test sequence of the dual-task paradigm (primary baseline, secondary baseline, and dual-task testing) as well as the sequence of the listening conditions within the primary baseline and dual-task testing were randomized. An identical test sequence was used for matched participants. After two weeks (mean interval: 14.8 days, range: 9 – 23 days), all participants re-executed the dual-task paradigm using an identical test sequence and the same prioritization instructions.

### 2.4 Statistical Analysis

Statistical analysis was performed using SPSS Version 29. A significance level of 0.01 was applied to all statistical tests to compensate for multiple comparisons. First, descriptive statistics were calculated for the participants’ characteristics (e.g., age, gender, educational level, and hearing sensitivity) and dual-task parameters. Second, Wilcoxon Singed Rank tests were conducted to compare scores on the primary speech-understanding task between the baseline and dual-task conditions across all listening conditions and test sessions. This analysis evaluated the assumption that participants’ performance on the primary task remained stable between baseline and dual-task conditions at the group level, as has been standardly performed in previous research (e.g., Degeest et al., 2022). Finally, to gain deeper insight into individual-level prioritization strategy, patterns of dual-task interference were calculated. For these categorical variables, Fisher’s exact tests were used to assess whether there was an association between interference patterns and group (i.e., non-prioritizing and prioritizing group) across all listening conditions and test sessions.

## 3. RESULTS

### 3.1 Participants

Out of the 48 participants involved, one exhibited hearing loss. Consequently, this participant and his/her/them matching person were excluded, resulting in a total of 46 included participants. Table 1 shows the demographic data of the non-prioritizing and prioritizing group which were matched for gender, age, and educational level.

**Table 1:** Demographic data of the non-prioritizing (NP) and prioritizing (P) group.

|  |  | NP | P |
| --- | --- | --- | --- |
| Gender (n, %) | Male | 9 (39.13%) | 9 (39.13%) |
|  | Female | 14 (60.87%) | 14 (60.87%) |
| Age (years) | Mean (SD) | 36.74 (13.92) | 36.79 (13.66) |
|  | Range | 18.08 – 59.50 | 18.83 – 59.09 |
| Educational level (n, %) | ≤ 12 years | 3 (13.04%) | 3 (13.04%) |
|  | > 12 years | 20 (86.96%) | 20 (86.96%) |
| Hearing sensitivity*<br>right ear (dB HL) | Mean (SD) | 5.05 (4.98) | 4.24 (5.51) |
|  | Range | -2.00 – 16.00 | -2.00 – 16.00 |
| Hearing sensitivity*<br>left ear (dB HL) | Mean (SD) | 4.13 (4.51) | 4.13 (5.95) |
|  | Range | -5.00 – 10.00 | -5.00 – 19.00 |

\* the audiometric threshold averaged over 0.5, 1, 2, and 4 kHz

### 3.2 Group-level performance stability on the primary task

The primary speech understanding scores at baseline and dual-task conditions for the non-prioritizing and prioritizing groups are presented in Table 2 for both test sessions. The Wilcoxon Signed Rank tests revealed no statistically significant differences (p > 0.01) between baseline and dual-task primary speech understanding scores across all listening conditions and test sessions, with one exception. A significant difference was observed for the +8 dB SNR listening condition in the non-prioritization group during test session 1 (p = 0.003), indicating a deviation from group-level performance stability in this specific case. Apart from this isolated condition, participants’ primary task performance remained stable between baseline and dual-task conditions at the group level.

**Table 2:** Primary speech understanding scores at baseline and dual-task conditions for the non-prioritizing (NP) and prioritizing (P) group at test sessions 1 and 2.

| Test session | Group | Listening condition | Baseline |  |  | Dual-task |  |  | Wilcoxon Singed-Rank test |
| --- | --- | --- | --- | --- | --- | --- | --- | --- | --- |
|  |  |  | Mean ± SD | Median | Range | Mean ± SD | Median | Range |  |
| 1 | NP | Quiet | 12.00 ± 1.68 | 12.00 | 9.00 - 15.00 | 11.17 ± 2.27 | 11.00 | 5.00 - 15.00 | Z = -1.585, p = 0.113 |
|  |  | SNR+8 | 13.22 ± 1.44 | 13.00 | 10.00 - 15.00 | 12.35 ± 1.67 | 12.00 | 9.00 - 15.00 | Z = -2.947, p = 0.003* |
|  |  | SNR+4 | 13.39 ± 1.50 | 14.00 | 10.00 - 15.00 | 12.96 ± 1.74 | 13.00 | 9.00 - 15.00 | Z = -1.154, p = 0.249 |
|  |  | SNR0 | 12.30 ± 1.89 | 12.00 | 8.00 - 15.00 | 11.96 ± 1.92 | 11.00 | 9.00 - 15.00 | Z = -1.316, p = 0.188 |
|  |  | SNR-4 | 12.70 ± 1.77 | 13.00 | 9.00 - 15.00 | 12.74 ± 1.94 | 12.00 | 9.00 - 15.00 | Z = 0.000, p = 1.000 |
|  | P | Quiet | 12.48 ± 2.00 | 13.00 | 9.00 - 15.00 | 12.87 ± 2.05 | 14.00 | 8.00 - 15.00 | Z = -0.936, p = 0.349 |
|  |  | SNR+8 | 13.74 ± 1.45 | 14.00 | 11.00 - 15.00 | 13.35 ± 1.15 | 14.00 | 11.00 - 15.00 | Z = -1.699, p = 0.089 |
|  |  | SNR+4 | 13.74 ± 1.25 | 14.00 | 10.00 - 15.00 | 13.83 ± 1.34 | 14.00 | 10.00 - 15.00 | Z = -0.520, p = 0.603 |
|  |  | SNR0 | 12.48 ± 1.47 | 12.00 | 10.00 - 15.00 | 12.91 ± 1.08 | 13.00 | 11.00 - 15.00 | Z = -1.422, p = 0.155 |
|  |  | SNR-4 | 13.61 ± 1.44 | 14.00 | 10.00 - 15.00 | 13.35 ± 1.72 | 14.00 | 8.00 - 15.00 | Z = -0.522, p = 0.602 |
| 2 | NP | Quiet | 13.43 ± 1.31 | 13.00 | 11.00 - 15.00 | 12.52 ± 2.09 | 12.00 | 8.00 - 15.00 | Z = -2.429, p = 0.015 |
|  |  | SNR+8 | 13.48 ± 1.04 | 14.00 | 12.00 - 15.00 | 12.78 ± 1.68 | 13.00 | 9.00 - 15.00 | Z = -1.936, p = 0.053 |
|  |  | SNR+4 | 13.52 ± 1.08 | 14.00 | 12.00 - 15.00 | 13.52 ± 1.75 | 14.00 | 10.00 - 15.00 | Z = -0.112, p = 0.911 |
|  |  | SNR0 | 13.35 ± 1.30 | 13.00 | 10.00 - 15.00 | 12.65 ± 1.70 | 13.00 | 9.00 - 15.00 | Z = -1.917, p = 0.055 |
|  |  | SNR-4 | 14.26 ± 0.96 | 15.00 | 12.00 - 15.00 | 13.70 ± 1.18 | 13.00 | 12.00 - 15.00 | Z = -2.167, p = 0.030 |
|  | P | Quiet | 14.04 ± 0.98 | 14.00 | 12.00 - 15.00 | 13.48 ± 1.53 | 14.00 | 9.00 - 15.00 | Z = -1.904, p = 0.057 |
|  |  | SNR+8 | 14.00 ± 0.95 | 14.00 | 12.00 - 15.00 | 14.04 ± 0.98 | 14.00 | 12.00 - 15.00 | Z = -0.183, p = 0.855 |
|  |  | SNR+4 | 14.04 ± 1.02 | 14.00 | 11.00 - 15.00 | 14.13 ± 0.87 | 14.00 | 12.00 - 15.00 | Z = -0.443, p = 0.658 |
|  |  | SNR0 | 13.78 ± 1.20 | 14.00 | 11.00 - 15.00 | 13.43 ± 1.38 | 14.00 | 11.00 - 15.00 | Z = -1.066, p = 0.286 |
|  |  | SNR-4 | 14.35 ± 0.71 | 14.00 | 13.00 - 15.00 | 13.87 ± 1.10 | 14.00 | 11.00 - 15.00 | Z = -2.066, p = 0.039 |

SNR: signal-to-noise ratio
SD: standard deviation
\* statistically significant, p < 0.01

### 3.3 Individual patterns of dual-task interference

The individual patterns of dual-task interference were assessed by plotting the DTE of the primary and secondary task against each other for both groups during the first (Figure 2) and second (Figure 3) test sessions. In these figures, each marker (circle or rhombus) represents an individual. Note that markers may overlap if multiple participants exhibited the same outcome. Table 3 displays the number of participants corresponding to each dual-task interference pattern across all listening conditions for test sessions 1 and 2.

**Figure 2:**
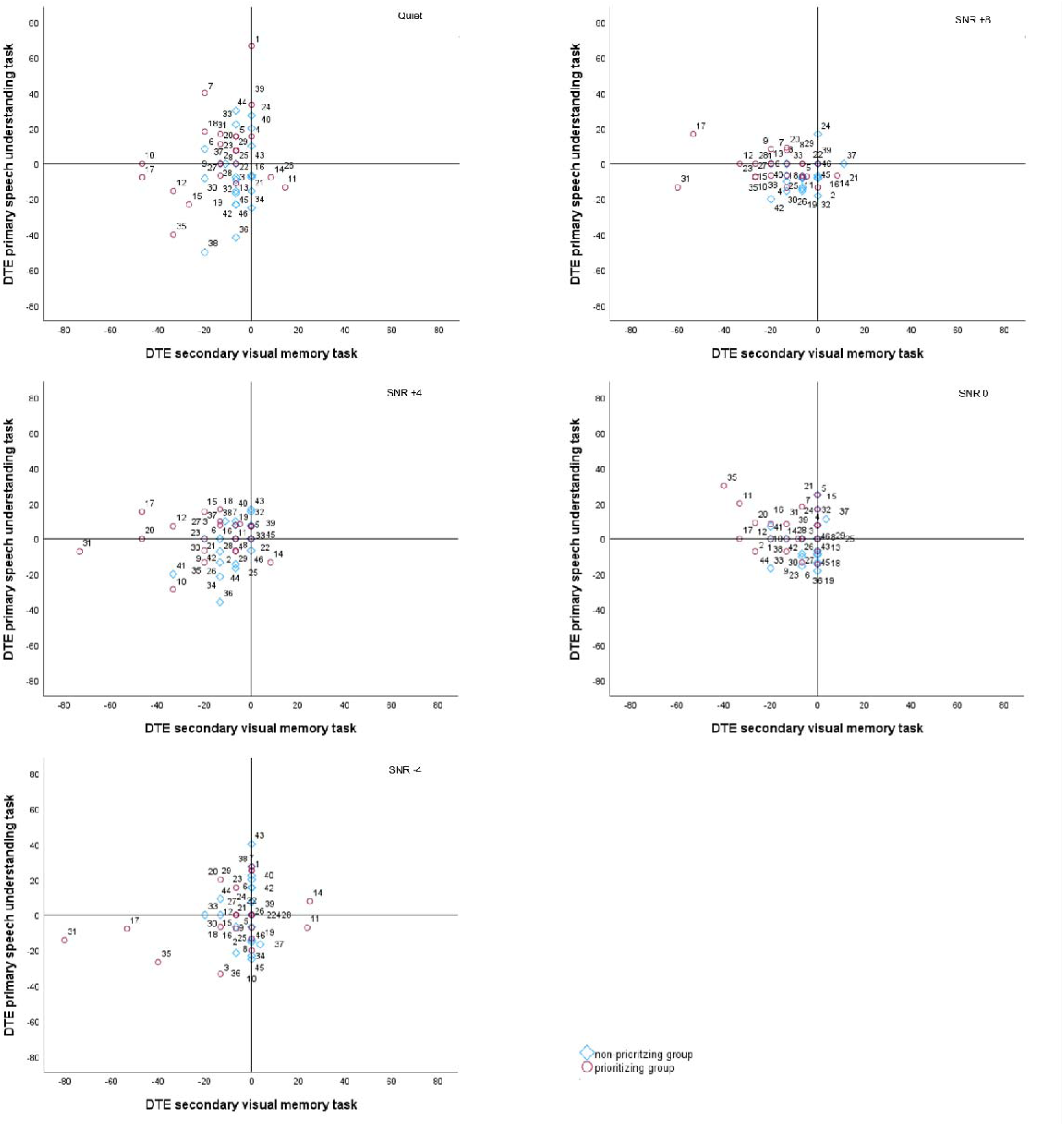
Framework of the dual-task interference patterns for the non-prioritizing and prioritizing groups at each listening conditions at test session 1.

**Figure 3:**
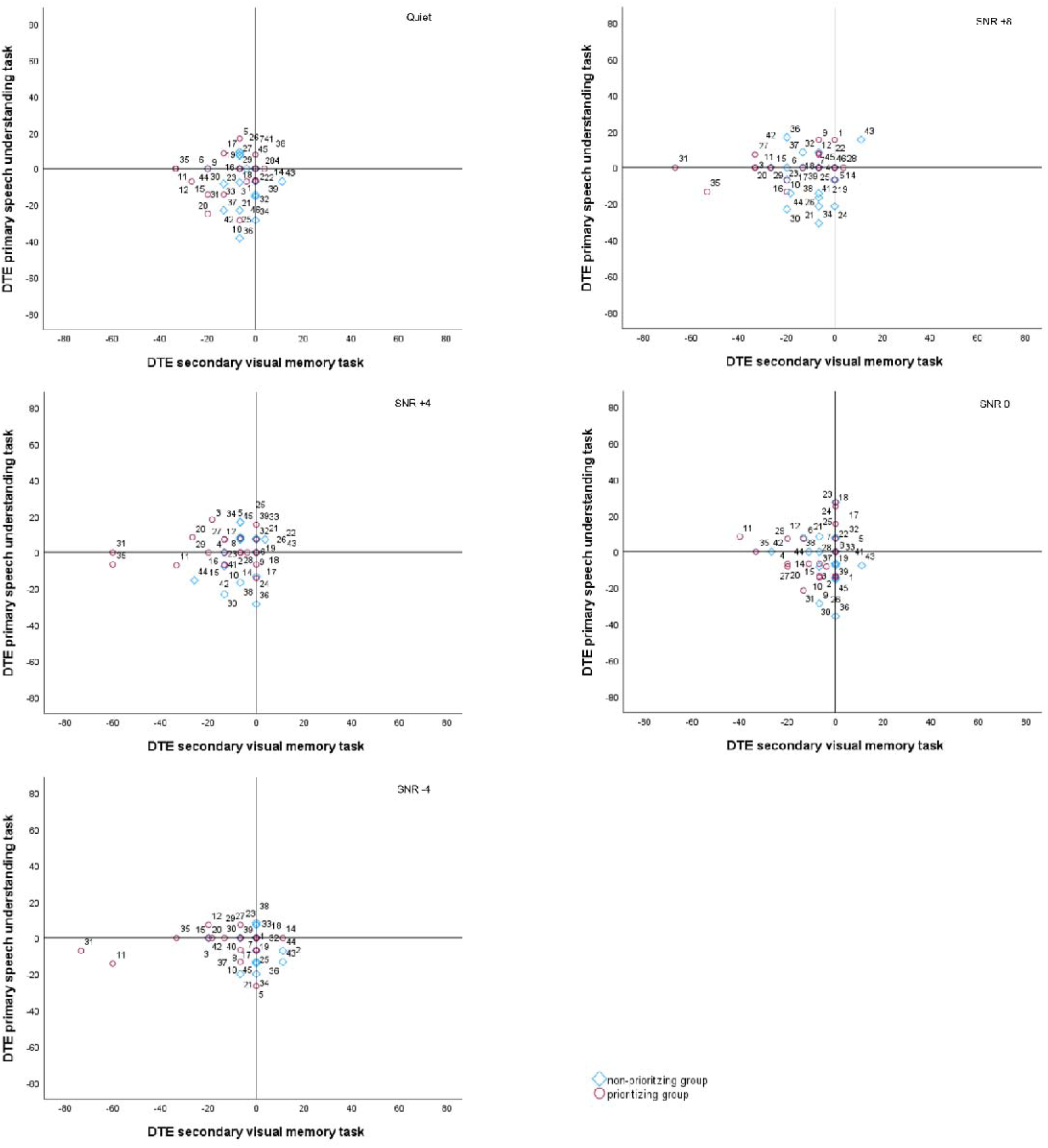
Framework of dual-task interference patterns for the non-prioritizing and prioritizing groups at each listening conditions at test session 2.

**Table 3:**
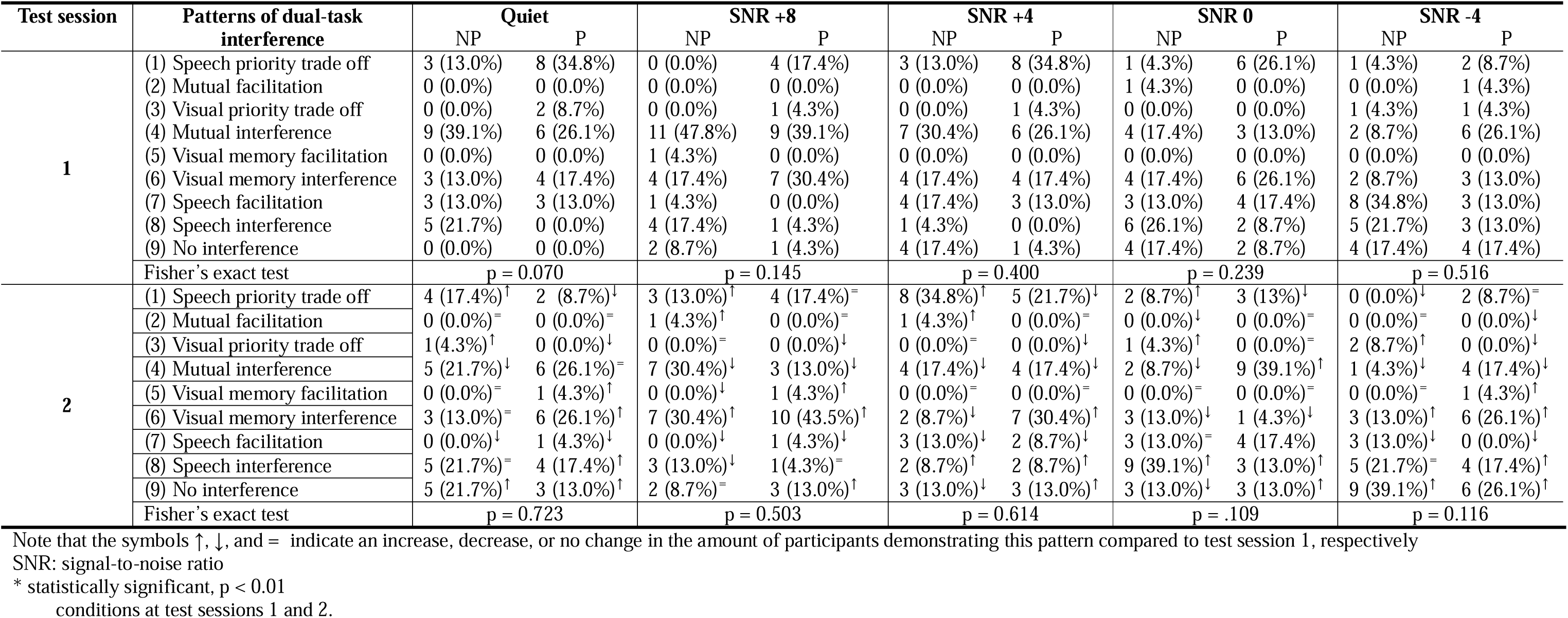
The number (n, %) of participants (NP = non-prioritizing group, P = prioritizing group) corresponding to each dual-task interference pattern across all listening conditions at test sessions 1 and 2.

For the first test session, the Fisher’s exact test showed no statistically significant association (p > 0.01) between the patterns of dual-task interference and the group to which the participants belong for none of the listening conditions (Table 3). Descriptive analysis revealed the application of diverse strategies for executing the dual-task. Overall, the non-prioritizing group employed mutual interference (pattern 4) and speech interference (pattern 8) more frequently than the prioritizing group. Conversely, the prioritizing group exhibited a higher occurrence of visual memory interference (pattern 6) and speech priority trade-off (pattern 1) compared to the non-prioritizing group. Notably, almost all participants of both the non-prioritizing and prioritizing groups used different strategies across the listening conditions within the first test session (Table 4). There was no clear or consistent pattern in strategy use, suggesting a lack of a definitive approach across conditions.

**Table 4:** The number of strategies (n,%) used across the five listening conditions by the participants (NP = non-prioritizing group, P = prioritizing group) for test sessions 1 and 2.

|  | Test session 1 |  | Test session 2 |  |
| --- | --- | --- | --- | --- |
|  | NP | P | NP | P |
| 1 strategy used | 1 (4.3%) | 0 (0.0%) | 0 (0.00%) | 2 (8.7%) |
| 2 strategies used | 4 (17.4%) | 7 (30.4%) | 5 (21.7%) | 6 (26.1%) |
| 3 strategies used | 9 (39.1%) | 9 (39.1%) | 11 (47.8%) | 8 (34.8%) |
| 4 strategies used | 8 (34.8%) | 7 (30.4%) | 6 (26.1%) | 5(21.7%) |
| 5 strategies used | 1 (4.3%) | 0 (0.0%) | 1 (4.3%) | 2 (8.7%) |

Regarding the second test session, the Fisher’s exact test showed no statistically significant association (p > 0.01) between the patterns of dual-task interference and the group to which the participants belong for none of the listening conditions (Table 3). When descriptively analysing the patterns of dual-task interference across the two test sessions, a shift in the pattern distribution was apparent (Table 3). In both the prioritizing and non-prioritizing group, a notable shift was observed, characterized by a decreased number of participants grappling with mutual interference (pattern 4) across nearly all listening conditions. Hence, participants showcased improved scores in at least one aspect of the dual-task condition during the second test session compared to the first test session. Furthermore, the prioritizing group tented to employ visual memory interference (pattern 6), speech interference (pattern 8), and no interference (pattern 9) more often during the second test session, while the non-prioritizing group leaned more towards speech priority trade-off (pattern 1) than during the first test session across nearly all listening conditions. As within the first test session, the majority of participants in both the prioritizing and non-prioritizing group employed varied strategies across the listening conditions during the second test session (Table 4).

## 4. DISCUSSION

Understanding how listeners execute a dual-task paradigm for listening effort would provide a benchmark for future studies and clinical implementations. Therefore, this study aimed to examine the prioritization strategy employed by individuals during a dual-task paradigm for listening effort.

Within the current study, the prioritizing group had more participants who achieved stable or better scores on the primary task in the dual-task condition compared to the non-prioritizing group. This observation might indicate an influence of instruction that does cause attentional and cognitive abilities to be more allocated to the primary task in the prioritizing group. In the study of Ceuleers et al. (2024), a trend was observed where more participants exhibited a speech priority trade-off pattern as listening conditions became more challenging. Ceuleers et al. suggested this could be due to the explicit instruction to prioritize the primary speech understanding task in the dual-task condition. In their baseline condition, where no such emphasis was placed on the primary task, participants may have put less effort into speech understanding compared to the dual-task condition. However, in the current study, there was considerable variability in the prioritizing strategy employed at the individual level across listening conditions and test sessions, regardless the given prioritization instructions. Hence, in line with previous research on children (Choi et al., 2008), providing prioritization instructions was insufficient to ensure that an individual would primarily focus on the primary task and consistently adhere to this strategy across listening conditions and test sessions.

This finding has important implications for the commonly used formula to evaluate listening effort in dual-task paradigms. Specifically, the DTE of the secondary task is traditionally used as a measure of listening effort, based on the assumption of stable primary task performance across baseline and dual-task conditions (Gagne et al., 2017). This assumption is typically assessed at the group level (e.g. Degeest et al., 2022). However, the current results demonstrated that group-level stability in primary task performance can obscure individual differences and fails to provide insights into the prioritization strategy employed when individuals’ cognitive capacity is exceeded. Furthermore, the instruction given to participants did not appear to guide or influence the strategy they choose. Instead, individuals seem to adopt a variety of strategies, with no clear or consistent approach emerging across individuals. This lack of a clear pattern highlights the complexity of individual decision-making and strategy use in the context of dual-tasking. Also, if only the DTE of the secondary task is taken into consideration, disregarded the DTE on the primary task, incomplete conclusion will be made. For example, when evaluating the impact of hearing rehabilitation on listening effort through the standard formula, no change in the dual-task cost of the secondary task would be interpreted as an indication that the hearing rehabilitation had no effect on listening effort. However, no change in the dual-task cost of the secondary task following rehabilitation may occur with a decrease in the dual-task cost of the primary task. This scenario implies that, due to rehabilitation, more attentional and cognitive resources were accessible which were directed towards the primary speech understanding task, suggesting a decline in listening effort (Gagne et al., 2017). This example emphasizes how an incomplete evaluation of the dual-task performance can offer misleading information.

While a dual-task paradigm for listening effort is widely used, the present study raised certain reservations about its applicability. First, the standard formula provides an insufficient or partial representation as it fails to account for dual-task interference. Second, evaluating patterns of dual-task interference may provide insight into the strategy used to perform a dual-task paradigm for listening effort but cannot serve as a real metric for listening effort. Last, there seems to be no singular strategy when performing a dual-task paradigm for listening effort, not even within the same individual. Therefore, as suggested by Kuchinsky et al. (2024), it is essential to critically evaluate current methodologies and consider revisiting foundational approaches to better integrate theoretical models of resource capacity, allocation, and task switching in dual-task paradigms for measuring listening effort.

### 4.1 Strengths, limitations and future perspectives

While the concept of integrating patterns of dual-task interference into research on listening effort was put forth years ago (Degeest et al., 2021; Gagne et al., 2017), this study is, to the best of our knowledge, one of the first to actually implement it. The results provided essential information on how normal-hearing listeners execute a dual-task paradigm for listening effort, questioning its applicability as a metric for listening effort.

Nevertheless, the current dual-task paradigm has limitations. Some participants exhibited no interference. This suggests that for both the primary and secondary tasks, there were no differences in performance between the baseline and dual-task conditions. For these participants, the dual-task may have been insufficiently challenging, implying that their cognitive capacity was not exceeded, thereby eliminating the necessity to prioritize between tasks. Also, a learning effect might be present since a reduced number of participants handling mutual interference was observed when comparing the performances on the first and second test sessions. Hence, some participants scored better on at least one aspect of the dual-task condition during the second test session. A limitation of using patterns of dual-task interference is that the data is categorical (nine distinct patterns), making it challenging to perform thorough statistical analyses, particularly because the number of participants was unevenly distributed across the patterns. The current study population does not represent the entire audiological target population. Hence, the current results can be considered a first exploration and should, therefore, be further investigated within diverse populations (such as individuals with hearing loss and/or hearing devices) as well as with diverse dual-task paradigms (concurrent and sequential, such as delayed recall). Apart from assessing the dual-task paradigm’s reliability as a metric of listening effort, the patterns of dual-task interference can be used as a counseling tool to enhance listeners’ comprehension of the limited capacity theory and the subsequent experienced listening effort.

### 4.2 Conclusion

Providing prioritization instructions when performing a dual-task paradigm for listening effort was insufficient to ensure that an individual will mainly focus on the primary task and consistently adhere to this strategy across listening conditions and test sessions. These results raised certain reservations about the current usage of dual-task paradigms for listening effort.

## Data Availability

All data produced in the present study are available upon reasonable request to the authors

## Acknowledgements

None to report

